# Assessing agreement between different polygenic risk scores in the UK Biobank

**DOI:** 10.1101/2022.02.09.22270719

**Authors:** Lei Clifton, Jennifer A Collister, Xiaonan Liu, Tom J Littlejohns, David J Hunter

**Affiliations:** Nuffield Department of Population Health, University of Oxford, Oxford, United Kingdom; Department of Epidemiology, Harvard TH Chan School of Public Health, Boston, MA, United States of America

## Abstract

Polygenic risk scores (PRS) are proposed to be used in clinical and research settings for risk stratification. However, there are limited investigations on how different PRS diverge from each other for risk prediction of individuals.

We compared two recently published PRS for each of three conditions, breast cancer, hypertension and dementia, to assess the stability of running these algorithms for risk prediction in a single large population. We used imputed genotyping data from the UK Biobank (UKB) prospective cohort, limited to the White British subset.

We found that:

1. Only 65%-79% of SNPs in the first PRS were represented in the more recent PRS for all three diseases, after having taken linkage disequilibrium (LD) into account (R^2^ >0.8).
2. Although the difference in the area under the received operator curve (AUC) obtained using the two PRS is hardly appreciable for all three diseases, there were large differences in individual risk prediction between the two PRS.

We found substantial discordance between different PRS for the same disease, indicating that individuals could receive different medical advice depending on which PRS is used to assess their genetic susceptibility. It is desirable to resolve this uncertainty before using PRS for risk stratification in clinical settings.

## 2. Introduction

Genome-wide association studies (GWAS) have revealed that the inherited genetic component of most traits not due to variations in a single gene is highly polygenic. Dozens or thousands of single nucleotide polymorphisms (SNPs) can be combined to produce a polygenic risk score (PRS) representing an individual’s genetic propensity for a given trait or disease.

There is much enthusiasm for the use of PRS to inform individuals about their risk of future health conditions, either as stand-alone information, or combined with non-genetic data in integrated risk scores [1], [2]. PRS have been proposed in a wide variety of settings such as prioritizing people for disease screening, informing the prescription of preventive medicines, and even in embryo selection [3]. Variations in single genes associated with diseases are already utilised in a clinical setting, however, recent large-scale studies have found that PRS could potentially identify a greater proportion of at risk individuals [4].

Early in the development of PRS, researchers [5], [6] quantified the degree to which a PRS with a limited number of SNPs would misclassify people, when compared with future PRS with many additional variants. At the time it was thought that once dozens, or even hundreds of SNPs were included, diminishing returns would set in and the PRS would be relatively stable. This perception appeared to be supported by the fact that in many large multi-study consortia, additional SNPs now being identified have very small odds ratios (OR; as low as 1.02 per allele or less) and the area under the curve (AUC) for Receiver Operator Characteristic (ROC) curves of newer versions of the PRS are only minimally higher than that of previous versions.

Recent discussion on the use of PRS in the clinic has largely focused on the reporting standards for the derivation and archiving of PRS [7], [8], the health economic value of PRS, the potential contribution of PRS to health disparities given the limited databases available for non-European ancestry populations [9], the most appropriate way for benefiting the patients [10], [11], and the means of communicating PRS to patients or members of the public.

During the construction of PRS, there are multiple design options for deciding the number of SNPs to include and for assigning an appropriate weight to each SNP. Consequently, multiple sets of SNPs exist, resulting in multiple PRS for the same trait. For example, the 313-SNP PRS for breast cancer [12] was developed using hard-thresholding stepwise forward regression, whereas the 118k-SNP PRS [13] was selected by the penalised regression “lassosum” and the highest pseudo-R^2^. However, these PRS are typically compared at a population level using metrics such as the AUC or OR, and limited attention has been paid to how they differ from each other for risk prediction of individuals.

## 3. Materials and Methods

### 3.1 Study populations

We used the genetic data from the UK Biobank (UKB), a large-scale population-based prospective cohort study of approximately 500,000 individuals aged 40-69 years at recruitment across the United Kingdom between March 2006 and October 2010. The ethical collection of sample and full details of the genotyping and imputation are described elsewhere [14], [15].

Our study populations for each of the three disease outcomes are defined as follows:

- The breast cancer eligible population was women who had not had breast cancer, carcinoma in situ or mastectomy prior to baseline.
- For the hypertension eligible population, we excluded individuals with missing or implausible systolic blood pressure (SBP) measurements (< 70 or > 270 mmHg) at baseline, and those with prior Major Adverse Cardiovascular Events (MACE).
- The dementia eligible population was restricted to individuals without prior diagnosis of dementia or Alzheimer’s disease.

Disease ascertainment of UKB during the following-up period utilised linkage to death registry, cancer registry, and Hospital Episode Statistics (HES). Hypertension is defined as SBP>=140 at baseline; the International Classification of Diseases (ICD) code for breast cancer and dementia can be found in Supplementary Tables 13-15.

### 3.2 Calculating PRS

We computed PRS of an individual *j* by the weighted sum of trait-associated SNPs,

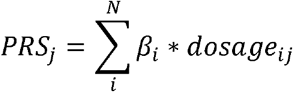

where N is the total number of SNPs, *β*_*i*_ is the effect size (or beta) of SNP *i*, and *dosage*_*ij*_ is the number of effect alleles (usually encoded as 0, 1 or 2 in SNP *i* for individual *j* for the effect allele).

We used published effect size of SNPs, and applied genetic quality control (QC) pipelines for both SNPs and samples. During SNP QC, we removed ambiguous SNPs (A/T or C/G SNPs with MAF > 0.49) and rare variants with MAF < 0.005; we only retained SNPs with high imputation quality (imputation information score > 0.4) (Supplementary Table 1). During sample QC, we excluded participants who were sex-discordant, outliers for missingness or heterozygosity, or related at 3^rd^ degree or higher, using UKB Data Field 22020.

**Table 1:** PRS compared for each outcome and their performance characteristics in the UK Biobank. N: number of participants whose PRS score was obtained. nSNPs: number of SNPs in PRS prior to genetic quality control. OR: odds ratio for top 1% vs middle quintile of PRS from multivariable logistic regression model adjusted for age, sex, genotyping array and first 5 PCs. AUC: area under receiver-operating curve using predicted risk from multivariable logistic regression model containing age, sex, continuous PRS, genotyping array and first 5 PCs. NRI: continuous net reclassification index using predicted risks from two multivariable logistic regression models containing age, sex, continuous PRS for this disease, genotyping array and first 5 PCs. The model containing PRS-B is considered the “updated” model. *r*: Pearson correlation coefficient between the two continuous PRS for this disease. LD: number (%) of SNPs in PRS-A which either appear in or are in linkage disequilbrium (R^2^ > 0.8) with SNPs in PRS-B. Breast cancer models are not adjusted for sex because its population is restricted to females.

| Disease (N) | PRS | nSNPs | OR (95% CI) | AUC (95% CI) | NRI (95% CI) | r | LD: $R^2 > 0.8$ (%) overlap) |
| --- | --- | --- | --- | --- | --- | --- | --- |
| Breast cancer (171,490) | A | 313 | 3.41<br>(2.89, 4.03) | 0.64<br>(0.63, 0.64) | 0.03<br>(0.00, 0.05) | 0.65 | 225 (72%) |
|  | B | 118388 | 3.94<br>(3.36, 4.61) | 0.64<br>(0.63, 0.65) |  |  |  |
| Hypertension (317,581) | A | 267 | 1.83<br>(1.70, 1.98) | 0.69<br>(0.69, 0.69) | 0.16<br>(0.16, 0.17) | 0.66 | 210 (79%) |
|  | B | 884 | 2.18<br>(2.02, 2.35) | 0.70<br>(0.70, 0.70) |  |  |  |
| Dementia (335,689) | A | 20 | 1.78<br>(1.40, 2.26) | 0.80<br>(0.79, 0.80) | 0.09<br>(0.06, 0.12) | 0.73 | 13 (65%) |
|  | B | 39 | 2.30<br>(1.85, 2.87) | 0.80<br>(0.79, 0.81) |  |  |  |

For each of the three disease outcomes, we selected a pair of recently published PRS within two years of each other for each disease, where the sample sizes for PRS derivation are large. Our PRS for breast cancer and dementia are from the polygenic score (PGS) Catalogue (www.PGSCatalog.org) [16], while PRS for hypertension are from the literature [17], [18]. The earlier PRS is denoted as PRS-A (typically contains fewer SNPs), while the more recent one is PRS-B with more SNPs. For each pair of PRS, we compared the total number of SNPs and the overlap between scores by checking the number of SNPs in common between the scores or in high LD (R^2^>0.8).

We then computed PRS for those within the defined study population of each condition, restricting to genetically White British individuals using UKB Data Field 22006. This yields the final size N of each study population.

- For breast cancer, our “baseline” PRS-A (313 SNPs, PGS ID: PGS000004) [12] has been widely validated and is included in the current implementation of the BOADICEA breast cancer risk model [19], [20]. For the “comparison” PRS-B, we used a score containing (118,388 SNPs, PGS ID: PGS000511) [13] which was largely developed from the same Breast Cancer Association Consortium (BCAC) GWAS data as PRS-A [21].
- For hypertension, we selected PRS for SBP from the literature. SBP PRS-A contains 267 SNPs [22], which we compared to a later SBP PRS-B containing 884 SNPs [23] with effect sizes from the International Consortium of Blood Pressure-Genome Wide Association Studies (ICBP), Million Veteran Program (MVP) and Estonian Genomic Centre of the University of Tartu (EGCUT).
- PRS-A for dementia originally contained 22 SNPs (PGS ID: PGS000334) [24]; we subsequently removed the two APOE SNPs (rs429358 and rs7412) to avoid the APOE genotype dominating the PRS, leaving us with 20 SNPs. Our 39-SNP PRS-B [25] (PGS ID: PGS001775) used effect sizes from the International Genomics of Alzheimer’s Project (IGAP) GWAS [26]; we retained all the 39 SNPs since they do not include the two APOE SNPs.

### 3.3 Quantifying the stability of PRS

In each disease-specific study population, we calculated the correlation coefficient between each pair of PRS and the age- and sex-adjusted odds ratios (ORs) between various cut-points compared with the middle quintile of the PRS distribution. We then used predictions from a multivariable logistic regression model containing age, sex, the continuous PRS, genetic array, and first 5 PCs to compute the area under curve (AUC) for each PRS (Table 1). The continuous Net Reclassification Index (NRI) was used to compare PRS-A with PRS-B in multivariable logistic models (Table 1), whereas the categorical NRI was used in cross-classification of PRS percentile risk categories (Percentage reclassification for participants who experienced the outcome are shown in Supplementary Tables 4-6 and 10-12, for top 1% and top 5% risk categorisations, respectively).

## 4. Results

Our study populations were N=171,490 (cases=6,347) for breast cancer, N=317,581 (cases=137,649) for hypertension, and N=335,689 (cases=4,460) for dementia. For comparing different PRS, we focused on two aspects: firstly, the consistency of the selected SNPs and performance metrics. Then we assessed the correlation between each pair of PRS, and the extent to which PRS-B gave the same predictions for individuals as PRS-A.

We found that only 65%-79% of SNPs in PRS-A were represented in PRS-B for all three diseases, after having taken linkage disequilibrium (LD) into account (R^2^>0.8). This is somewhat surprising, as one might expect a newer score (PRS-B) to incorporate most of the previously identified SNPs from PRS-A.

Table 1 presents the performance characteristics of each PRS against the corresponding disease outcome in UKB. In each case the more recent PRS-B was associated with a slightly higher OR than the earlier PRS-A. For example, the OR of breast cancer among women in the top 1% compared to those in the middle quintile was 3.41 for PRS-A and 3.94 for PRS-B. Their corresponding AUCs were only minimally different (0.638 vs 0.641), and the ROCs looked almost identical (**Error! Reference source not found.Figure 1**). Similar results were obtained for hypertension (PRS-A OR=1.83, AUC=0.69; PRS-B OR=2.18, AUC=0.70) and dementia (PRS-A OR=1.78, AUC=0.80; PRS-B OR=2.30, AUC=0.80).

**Figure 1:**
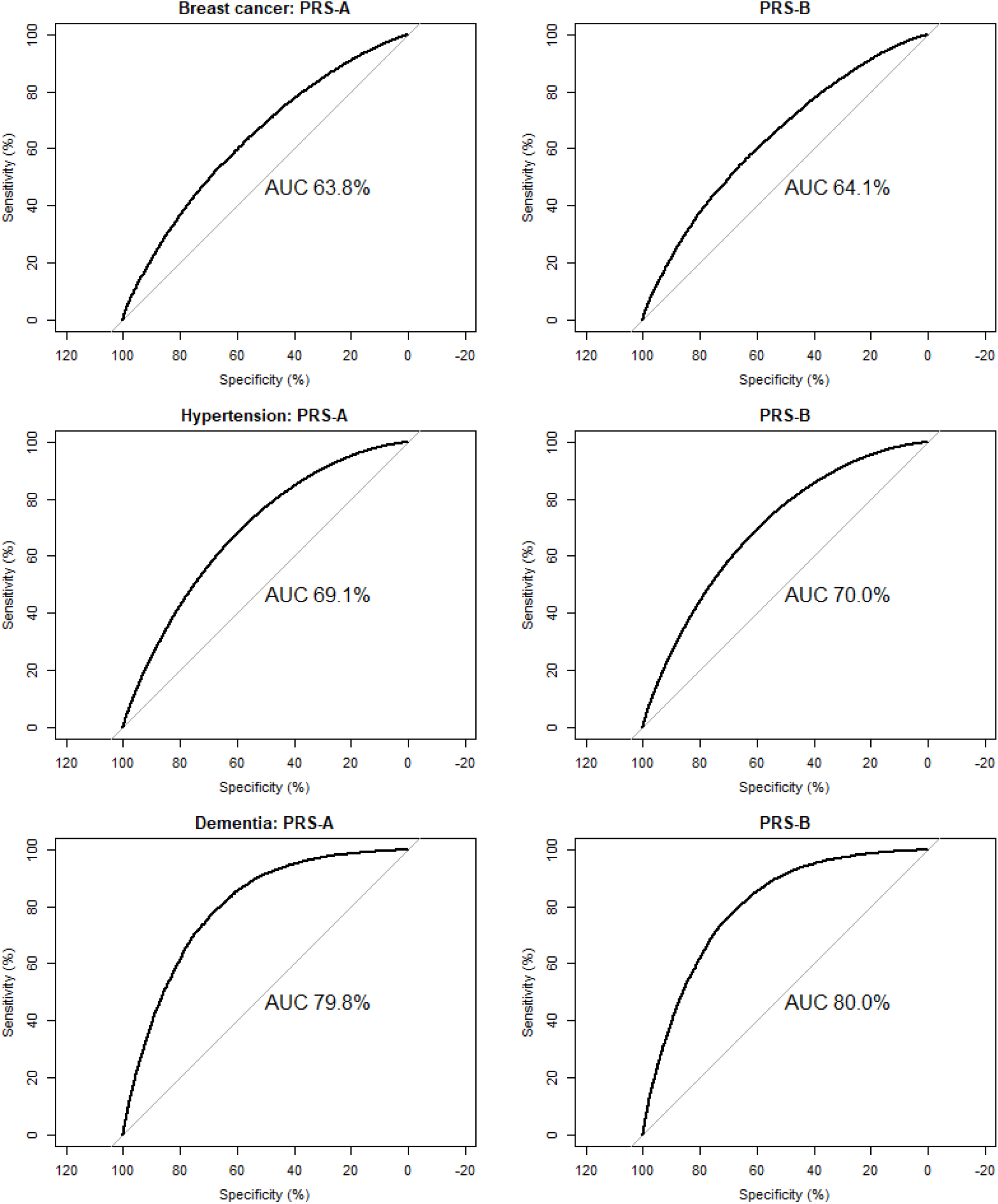
ROC plots obtained from predictions from multivariable logistic regression of age, sex, continuous PRS, genotyping array and first 5 PCs against disease outcome

Despite similar AUCs, PRS-A and PRS-B were not highly correlated for any outcome, with their Pearson correlation coefficient *r* only in the range of 0.65 to 0.73. Compatible with these correlation coefficients, there was substantial reclassification of predicted risk according to percentiles of PRS-A and PRS-B for all three diseases, as shown for breast cancer in Table 2. For women in the top 1% of breast cancer risk by PRS-A, only 23.1% were in the top 1% risk of PRS-B. The equivalent percentage was 22.9% and 22.7% for hypertension and dementia, respectively (Supplementary Tables 2-3). We focused on the top 1% of risk because of the widely-promulgated concept that these risks approximate those of the risks for monogenic traits [4].

**Table 2:** Cross-classification of predicted risk of breast cancer, according to the percentiles of each PRS. Number of participants are shown as n (col%, row%, cell%). Higher percentiles of PRS indicate increased risk of breast cancer; “≥ 99%” percentile corresponds to the top 1% risk.

| Percentiles of PRS-A | Percentiles of PRS-B |  |  |  |  |  |  |
| --- | --- | --- | --- | --- | --- | --- | --- |
|  | < 1% | 1-20% | 20-40% | 40-60% | 60-80% | 80-99% | ≥ 99% |
| < 1% | 345<br>(20.1,<br>20.1,<br>0.2) | 1140<br>(66.5,<br>3.5,<br>0.7) | 176<br>(10.3,<br>0.5,<br>0.1) | 44<br>(2.6,<br>0.1,<br>0.0) | 9<br>(0.5,<br>0.0,<br>0.0) | 1<br>(0.1,<br>0.0,<br>0.0) | 0<br>(0.0,<br>0.0,<br>0.0) |
| 1-20% | 1117<br>(3.4,<br>65.1,<br>0.7) | 15409<br>(47.3,<br>47.3,<br>9.0) | 8818<br>(27.1,<br>25.7,<br>5.1) | 4696<br>(14.4,<br>13.7,<br>2.7) | 2052<br>(6.3,<br>6.0,<br>1.2) | 490<br>(1.5,<br>1.5,<br>0.3) | 1<br>(0.0,<br>0.1,<br>0.0) |
| 20-40% | 198<br>(0.6,<br>11.5,<br>0.1) | 8788<br>(25.6,<br>27.0,<br>5.1) | 9989<br>(29.1,<br>29.1,<br>5.8) | 8053<br>(23.5,<br>23.5,<br>4.7) | 5210<br>(15.2,<br>15.2,<br>3.0) | 2051<br>(6.0,<br>6.3,<br>1.2) | 9<br>(0.0,<br>0.5,<br>0.0) |
| 40-60% | 43<br>(0.1,<br>2.5,<br>0.0) | 4648<br>(13.6,<br>14.3,<br>2.7) | 7998<br>(23.3,<br>23.3,<br>4.7) | 8908<br>(26.0,<br>26.0,<br>5.2) | 8050<br>(23.5,<br>23.5,<br>4.7) | 4607<br>(13.4,<br>14.1,<br>2.7) | 44<br>(0.1,<br>2.6,<br>0.0) |
| 60-80% | 11<br>(0.0,<br>0.6,<br>0.0) | 2098<br>(6.1,<br>6.4,<br>1.2) | 5296<br>(15.4,<br>15.4,<br>3.1) | 7988<br>(23.3,<br>23.3,<br>4.7) | 10047<br>(29.3,<br>29.3,<br>5.9) | 8688<br>(25.3,<br>26.7,<br>5.1) | 170<br>(0.5,<br>9.9,<br>0.1) |
| 80-99% | 1<br>(0.0,<br>0.1,<br>0.0) | 497<br>(1.5,<br>1.5,<br>0.3) | 2006<br>(6.2,<br>5.8,<br>1.2) | 4574<br>(14.0,<br>13.3,<br>2.7) | 8736<br>(26.8,<br>25.5,<br>5.1) | 15674<br>(48.1,<br>48.1,<br>9.1) | 1095<br>(3.4,<br>63.8,<br>0.6) |
| ≥ 99% | 0<br>(0.0,<br>0.0,<br>0.0) | 3<br>(0.2,<br>0.0,<br>0.0) | 15<br>(0.9,<br>0.0,<br>0.0) | 35<br>(2.0,<br>0.1,<br>0.0) | 194<br>(11.3,<br>0.6,<br>0.1) | 1072<br>(62.5,<br>3.3,<br>0.6) | 396<br>(23.1,<br>23.1,<br>0.2) |

Using a more relaxed risk category, participants in the top 5% of risk for breast cancer by PRS-A, only 35.7% were in the top 5% risk by PRS-B. The equivalent percentage was 35.8% and 40.0% for hypertension and dementia, respectively (Supplementary Tables 7-9).

## 5. Discussion

The clinical utility of PRS depends on the clinical validity of the predictions. Clinical validity is not only dependent on the information PRS give on the risk of future events, but also on the stability of these estimates. Here we show that for three common conditions (breast cancer, hypertension and dementia), the risk estimates derived from different PRS would result in very different information being given to a high proportion of people. Choice of the PRS may also influence the use of PRS as covariates or effect modifiers in epidemiologic analyses.

We found that the more recent PRS-B had minimal increase in AUC compared to the older PRS-A, in line with the small improvement measured by net reclassification index (NRI). However, the PRS differed substantially in how they assigned participants into risk categories with a substantial proportion of individuals classified at high risk by one PRS, not so classified by the other PRS. This suggests a major potential problem for the use of these PRS in clinical practice, given the changes in clinical recommendations associated with labelling a person in the same category of risk as a monogenic disorder. Our results demonstrated large differences across all percentiles of risk; although the clinical consequences at the lower percentiles may not be as extreme as at the higher percentiles, the clinical utility will still be reduced by incorrect classification.

While UKB data was used in the development of some of these PRS, since our interest is in comparing the classification of individuals by each score and not in developing a prediction model, we expect this to have little impact on the results.

We note that we have not established the reasons for the extent of misclassification between different PRS. It does not appear to be attributable solely to the number of SNPs included in the PRS – we show this for PRS comprised of over 100 thousand versus several hundred SNPs (breast cancer), for PRS composed of hundreds of SNPs (systolic blood pressure), and for PRS composed of <50 SNPs (dementia). The surprisingly small number of SNPs held in common by different PRS for the same condition published only a year apart indicates that different analytical methods used to derive the PRS may account for some of the discrepancies in classification, but understanding this phenomenon is clearly important for PRS selection in broad clinical practice. The correlations we observed between PRS for the same condition are in the same range as is seen for variation in risk predictors measured several years apart such as blood pressure and serum cholesterol, far from the “fixed” or “one-time” value at birth that is often assumed for PRS.

We are not the first to notice this phenomenon. For instance, Läll [27] compared the performance of four PRS in breast cancer prediction, noted that some of the correlations between them were as low as *r*=0.3, and observed that a “metaGRS” of the PRS performed better than any of the individual PRS [28]. However, the issue does not seem to be widely appreciated, and most publications comparing a new PRS with previous versions assert the superiority of the new PRS and do not address the issue of misclassification of risk between PRS. Our observation shows two PRS that only minimally differ in predictive performance on a population level may substantially differ in terms of individual risk classification, even among individuals with the same continental ancestry. This issue requires careful consideration before utilising PRS in real-world settings, because such an arbitrary element in health care is obviously undesirable. A person’s genetic profile is generally considered fixed at birth, leading to the widely held conviction that genetic susceptibility is an immutable value; however, our findings show that such assertions may be premature in the context of PRS which are still a developing area of research. While it is reasonable to expect incremental improvements in any risk prediction algorithm over time, these results suggest there is still considerable uncertainty associated with estimates of risk derived from different PRS for the same disease. It will be important to develop guidelines on best practice in constructing PRS to minimize the extent to which people will be given contradictory information over short periods of time.

## Data Availability

The data reported in this paper are available via application directly to the UK Biobank, https://www.ukbiobank.ac.uk

## 6. Declarations

### 6.1 Ethics approval and consent to participate

The UK Biobank study (*https://www.ukbiobank.ac.uk*) received ethical approval from the North West Multi-center Research Ethics Committee (REC reference: 11/NW/03820). All participants gave written informed consent before enrolment in the study, which was conducted in accordance with the principles of the Declaration of Helsinki. This study has been conducted under the UK Biobank application ID 33952.

### 6.2 Patient and community involvement

The analyses presented here are based on existing data from the UK Biobank cohort study, and the authors were not involved in participant recruitment. To the best of our knowledge, no patients were explicitly engaged in the design or implementation of the UK Biobank study. No patients were asked to advise on interpretation or writing these results. Results from UK Biobank are routinely disseminated to study participants via the study website and social media outlets.

### 6.3 Consent for publication

Yes.

### 6.4 Availability of data and material

Further summary data can be found in the Supplementary Materials; the authors are happy to provide further information upon the request of individual members of the public. Please note that the UK Biobank does not permit researchers to provide the raw data reported in this paper. However, interested readers are able to request the raw data via application directly to the UK Biobank *(https://www.ukbiobank.ac.uk)*.

### 6.5 Competing interests

The authors declare no competing interests. All authors declare no support from any organization for the submitted work, no financial relationship with any organization that might have an interest in the submitted work in the previous three years; no other relationship or activities that could appear to have influenced the submitted work.

### 6.6 Funding

The UK Biobank study was supported by the Wellcome Trust, Medical Research Council, Department of Health, Scottish government, and Northwest Regional Development Agency. It has also received funding from the Welsh Assembly government and British Heart Foundation. The analyses here were funded by the Cancer Research UK (grant no C16077/A29186), and supported by the Nuffield Department of Population Health, Oxford University.

### 6.7 Author contributions

Conceptualisation, investigation, supervision: LC, DJH;

Data curation, formal analysis, methodology, writing-original draft: LC, DJH, JAC, XL; Writing-review & editing: LC, DJH, JAC, XL, TJL.

All authors have revised the manuscript and agreed on its contents.

### 6.8 Transparency statement

The lead author affirms that this manuscript is an honest, accurate, and transparent account of the study being reported; that no important aspects of the study have been omitted; and that any discrepancies from the study as planned have been explained.

## 6.9 Acknowledgements

We thank the participants of the UK Biobank study for enabling us to conduct this research.

## Notes

### Competing Interest Statement

The authors have declared no competing interest.

### Summary of Updates

There was a typo in one of the author names in the previous version. This typo has been corrected in this version.

## References

[1] A. Torkamani, N. E. Wineinger, and E. J. Topol, “The personal and clinical utility of polygenic risk scores,” Nature Reviews Genetics, vol. 19, no. 9. Nat Rev Genet, pp. 581–590, 01-Sep-2018.

[2] T. Yanes, A. M. McInerney-Leo, M. H. Law, and S. Cummings, “The emerging field of polygenic risk scores and perspective for use in clinical care,” Human Molecular Genetics, vol. 29, no. R2. Hum Mol Genet, pp. R165–R176, 2020.

[3] L. C. A. M. Tellier, J. Eccles, N. R. Treff, L. Lello, S. Fishel, and S. Hsu, “Embryo screening for polygenic disease risk: Recent advances and ethical considerations,” Genes (Basel)., vol. 12, no. 8, Jul. 2021.

[4] A. V. Khera et al., “Genome-wide polygenic scores for common diseases identify individuals with risk equivalent to monogenic mutations,” Nature Genetics, vol. 50, no. 9. Nature Publishing Group, pp. 1219–1224, 01-Sep-2018.

[5] P. Kraft and D. J. Hunter, “Genetic Risk Prediction — Are We There Yet?,” N. Engl. J. Med., vol. 360, no. 17, pp. 1701–1703, Apr. 2009.

[6] N. Chatterjee, B. Wheeler, J. Sampson, P. Hartge, S. J. Chanock, and J. H. Park, “Projecting the performance of risk prediction based on polygenic analyses of genome-wide association studies,” Nat. Genet., vol. 45, no. 4, pp. 400–405, Apr. 2013.

[7] H. Wand, S. A. Lambert, C. Tamburro, and M. A. Iacocca, “Improving reporting standards for polygenic scores in risk prediction studies,” Nature, vol. 591, no. April 2020, 2021.

[8] Y. Ding et al., “Large uncertainty in individual polygenic risk score estimation impacts PRS-based risk stratification,” Nat. Genet., vol. 54, no. 1, pp. 30–39, Dec. 2021.

[9] C. M. Lewis and E. Vassos, “Polygenic risk scores: From research tools to clinical instruments,” Genome Med., vol. 12, no. 1, pp. 1–11, 2020.

[10] J. W. Knowles and E. A. Ashley, “Cardiovascular disease: The rise of the genetic risk score,” PLoS Med., vol. 15, no. 3, p. e1002546, Mar. 2018.

[11] C. M. Lewis and E. Vassos, Polygenic risk scores: From research tools to clinical instruments, vol. 12, no. 1. BioMed Central, 2020, pp. 1–11.

[12] N. Mavaddat et al., “Polygenic Risk Scores for Prediction of Breast Cancer and Breast Cancer Subtypes,” Am. J. Hum. Genet., vol. 104, no. 1, pp. 21–34, Jan. 2019.

[13] L. G. Fritsche et al., “Cancer PRSweb: An Online Repository with Polygenic Risk Scores for Major Cancer Traits and Their Evaluation in Two Independent Biobanks,” Am. J. Hum. Genet., vol. 107, no. 5, pp. 815–836, Nov. 2020.

[14] C. Bycroft et al., “The UK Biobank resource with deep phenotyping and genomic data,” Nature, vol. 562, no. 7726, pp. 203–209, 2018.

[15] R. Collins, “What makes UK Biobank special?,” The Lancet, vol. 379, no. 9822. pp. 1173–1174, Mar-2012.

[16] S. A. Lambert et al., “The Polygenic Score Catalog as an open database for reproducibility and systematic evaluation,” Nature Genetics, vol. 53, no. 4. Nature Publishing Group, pp. 420–425, 10-Mar-2021.

[17] H. R. Warren et al., “Genome-wide association analysis identifies novel blood pressure loci and offers biological insights into cardiovascular risk,” Nat. Genet., vol. 49, no. 3, pp. 403–415, Mar. 2017.

[18] E. Evangelou et al., “Genetic analysis of over 1 million people identifies 535 new loci associated with blood pressure traits,” Nat. Genet., vol. 50, no. 10, pp. 1412–1425, Oct. 2018.

[19] A. Lee et al., “BOADICEA: a comprehensive breast cancer risk prediction model incorporating genetic and nongenetic risk factors,” Genet. Med., vol. 21, no. 8, 2019.

[20] I. M. M. Lakeman et al., “Validation of the BOADICEA model and a 313-variant polygenic risk score for breast cancer risk prediction in a Dutch prospective cohort,” Genet. Med. 2020 2211, vol. 22, no. 11, pp. 1803–1811, Jul. 2020.

[21] K. Michailidou et al., “Association analysis identifies 65 new breast cancer risk loci,” Nat. 2017 5517678, vol. 551, no. 7678, pp. 92–94, Oct. 2017.

[22] H. R. Warren et al., “Genome-wide association analysis identifies novel blood pressure loci and offers biological insights into cardiovascular risk,” Nat. Genet. 2017 493, vol. 49, no. 3, pp. 403–415, Jan. 2017.

[23] E. Evangelou et al., “Genetic analysis of over 1 million people identifies 535 new loci associated with blood pressure traits,” Nat. Genet., vol. 50, no. 10, pp. 1412–1425, Oct. 2018.

[24] Q. Zhang et al., “Risk prediction of late-onset Alzheimer’s disease implies an oligogenic architecture,” Nat. Commun. 2020 111, vol. 11, no. 1, pp. 1–11, Sep. 2020.

[25] J. L. Ebenau et al., “Risk of dementia in APOE ε4 carriers is mitigated by a polygenic risk score,” Alzheimer’s Dement. Diagnosis, Assess. Dis. Monit., vol. 13, no. 1, Jan. 2021.

[26] B. W. Kunkle et al., “Genetic meta-analysis of diagnosed Alzheimer’s disease identifies new risk loci and implicates Aβ, tau, immunity and lipid processing,” Nat. Genet. 2019 513, vol. 51, no. 3, pp. 414–430, Feb. 2019.

[27] K. Läll et al., “Polygenic prediction of breast cancer: Comparison of genetic predictors and implications for risk stratification,” BMC Cancer, vol. 19, no. 1, Jun. 2019.

[28] M. Inouye et al., “Genomic Risk Prediction of Coronary Artery Disease in 480,000 Adults: Implications for Primary Prevention,” J. Am. Coll. Cardiol., vol. 72, no. 16, pp. 1883–1893, Oct. 2018.

