## Supplementary Tables for "Assessing agreement between different polygenic risk scores in the UK Biobank"

Lei Clifton^1*^, Jennifer A Collister^1^, Xiaonan Liu^1^, Thomas J Littlejohns^1^, David J Hunter^1,2^

1. Nuffield Department of Population Health, University of Oxford, Oxford, United Kingdom

2. Department of Epidemiology, Harvard TH Chan School of Public Health, Boston, MA, United States of America

* Corresponding Author

ORCID: 0000-0001-5595-8468

### 2 Table of PRS characteristics

Supplementary Table 1: PRS characteristics. The “Derivation dataset” columns describe, to the best of our knowledge, the source GWAS(s) from which the SNP effect sizes for each PRS were derived, for more information please see the source publication. The “Validation dataset” column describes which datasets the derived score was evaluated in within the source publication.

Abbreviations: BCAC: Breast Cancer Association Consortium, GWAS: Genome-Wide Association Study, MGI: Michigan Genomics Initiative, SBP: Systolic Blood Pressure, ICBP: International Consortium of Blood Pressure, IGAP: International Genomics of Alzheimer’s Project, AIBL: Australian Imaging, Biomarker & Lifestyle Flagship Study, Sydney MAS: Sydney Memory and Ageing Study, SCIENCe project: Subjective Cognitive Impairment Cohort.

| Disease | PRS | nSNPs | Source  (PGS ID) | Trait | Construction | Derivation dataset | | | Validation Dataset |
| --- | --- | --- | --- | --- | --- | --- | --- | --- | --- |
|  |  |  |  |  |  | Datasets | Cases | Ancestry |  |
| Breast cancer | A | 313 | [Mavaddat2019](https://doi.org/10.1016/j.ajhg.2018.11.002) ([PGS000004](https://www.pgscatalog.org/score/PGS000004/)) | Overall breast cancer | Hard-thresholding and stepwise forward regression, p<10^-5^ | [BCAC GWAS Summary Results: Breast Cancer Risk (2017)](https://bcac.ccge.medschl.cam.ac.uk/bcacdata/oncoarray/oncoarray-and-combined-summary-result/gwas-summary-results-breast-cancer-risk-2017/), described in Michailidou2017 | 94,075 cases and 75,017 controls | European | UK Biobank |
|  | B | 118,388 | [Fritsche2020](https://doi.org/10.1016/j.ajhg.2020.08.025) ([PGS000511](https://www.pgscatalog.org/score/PGS000511/)) | Overall breast cancer | Lassosum, s=0.5, lambda=0.004281 | [BCAC GWAS Summary Results: Breast Cancer Risk (2017)](https://bcac.ccge.medschl.cam.ac.uk/bcacdata/oncoarray/oncoarray-and-combined-summary-result/gwas-summary-results-breast-cancer-risk-2017/), described in Michailidou2017 | 94,075 cases and 75,017 controls | European | MGI |
| Hypertension | A | 267 | [Warren2017](https://doi.org/10.1038/ng.3768)  (Not in PGS Catalog) | SBP | Pairwise-independent, LD-filtered (r2 < 0.2) previously reported and novel variants | UK Biobank | 140,882 | European | Airwave |
|  | B | 884 | [Evangelou2018](https://doi.org/10.1038/s41588-018-0205-x)  ([PGS000812](https://www.pgscatalog.org/score/PGS000812/)) | SBP | Pairwise-independent, LD-filtered (r2 < 0.1) previously reported and novel variants | ICBP meta-analysis | 299,024 | European | UK Biobank |
| Dementia | A | 57 | [Najar2021](https://alz-journals.onlinelibrary.wiley.com/doi/10.1002/dad2.12142) ([PGS000812](https://www.pgscatalog.org/score/PGS000812/)) | Clinically defined Alzheimer’s Disease | p < 1e–5 | IGAP, described in [Kunkle2019](https://www.nature.com/articles/s41588-019-0358-2) | “*The final sample was 35,274 clinical and autopsy-documented Alzheimer’s disease cases and 59,163 controls.”* ^1^ | Non-Hispanic Whites | Gothenburg H70 Birth Studies and Prospective Population Study of Women |
|  | B | 39 | [Ebenau2021](https://doi.org/10.1002/dad2.12229) ([PGS001775](https://www.pgscatalog.org/score/PGS001775/)) | Alzheimer’s Disease | Genome-wide significant variants | IGAP, described in [Kunkle2019](https://www.nature.com/articles/s41588-019-0358-2), [Sims2017](https://www.nature.com/articles/ng.3916) | “*The final sample was 35,274 clinical and autopsy-documented Alzheimer’s disease cases and 59,163 controls.”* ^1^ | Non-Hispanic Whites | Amsterdam Dementia Cohort, SCIENCe project |

1.	Kunkle, B. W. *et al.* Genetic meta-analysis of diagnosed Alzheimer’s disease identifies new risk loci and implicates Aβ, tau, immunity and lipid processing. *Nat. Genet. 2019 513* **51**, 414–430 (2019).

### 3 Table of SNP QC

Supplementary Table 2: SNP QC

| Disease | PRS | nSNPs | Unavailable in UKB | Ambiguous | Imputation info < 0.4 | MAF < 0.005 | Remaining SNPs |
| --- | --- | --- | --- | --- | --- | --- | --- |
| Breast cancer | A | 313 | 7 | 0 | 0 | 1 | 305 |
|  | B | 118,388 | 0 | 43 | 107 | 2938 | 115,300 |
| Hypertension | A | 267 | 0 | 0 | 0 | 3 | 264 |
|  | B | 884 | 0 | 3 | 0 | 0 | 881 |
| Dementia | A | 57 | 0 | 0 | 0 | 0 | 57 |
|  | B | 39 | 0 | 0 | 0 | 1 | 38 |

### 4 Tables for top 1% risk

Supplementary Table 3: Cross-classification of predicted risk of hypertension among the whole study population (n=317,581), according to the percentiles of each PRS. Number of participants are shown as n (col%, row%, cell%). Higher percentiles of PRS indicate increased risk of hypertension; “≥ 99%” percentile corresponds to the top 1% risk.

| Percentiles of PRS A | Percentiles of PRS B | | | | | | |
| --- | --- | --- | --- | --- | --- | --- | --- |
|  | < 1% | 1-20% | 20-40% | 40-60% | 60-80% | 80-99% | ≥ 99% |
| < 1% | 758  (23.9, 23.9,  0.2) | 2024  (63.7,  3.4,  0.6) | 298  (9.4,  0.5,  0.1) | 78  (2.5,  0.1,  0.0) | 16  (0.5,  0.0,  0.0) | 2  (0.1,  0.0,  0.0) | 0  (0.0,  0.0,  0.0) |
| 1-20% | 2043  (3.4,  64.3,  0.6) | 29305  (48.6,  48.6,  9.2) | 15985  (26.5,  25.2,  5.0) | 8431  (14.0,  13.3,  2.7) | 3653  (6.1,  5.8,  1.2) | 923  (1.5,  1.5,  0.3) | 1  (0.0,  0.0, 0.0) |
| 20-40% | 290  (0.5,  9.1,  0.1) | 16014  (25.2,  26.5,  5.0) | 18776  (29.6,  29.6,  5.9) | 14854  (23.4,  23.4,  4.7) | 9954  (15.7,  15.7,  3.1) | 3617  (5.7,  6.0,  1.1) | 11  (0.0,  0.3,  0.0) |
| 40-60% | 68  (0.1,  2.1,  0.0) | 8391  (13.2,  13.9,  2.6) | 14996  (23.6, 23.6,  4.7) | 16595  (26.1,  26.1,  5.2) | 15006  (23.6,  23.6,  4.7) | 8381  (13.2,  13.9,  2.6) | 79  (0.1,  2.5,  0.0) |
| 60-80% | 14  (0.0,  0.4,  0.0) | 3664  (5.8,  6.1,  1.2) | 9803  (15.4,  15.4,  3.1) | 14988  (23.6,  23.6,  4.7) | 18520  (29.2, 29.2,  5.8) | 16226  (25.5,  26.9,  5.1) | 301  (0.5,  9.5, 0.1) |
| 80-99% | 3  (0.0, 0.1,  0.0) | 940  (1.6,  1.6,  0.3) | 3650  (6.0,  5.7,  1.1) | 8495  (14.1,  13.4,  2.7) | 16055  (26.6,  25.3,  5.1) | 29140  (48.3,  48.3,  9.2) | 2057  (3.4,  64.8,  0.6) |
| ≥ 99% | 0  (0.0, 0.0,  0.0) | 3  (0.1,  0.0,  0.0) | 10  (0.3, 0.0,  0.0) | 74  (2.3,  0.1,  0.0) | 311  (9.8,  0.5,  0.1) | 2051  (64.6,  3.4,  0.6) | 727  (22.9,  22.9,  0.2) |

Supplementary Table 4: Cross-classification of predicted risk of dementia among the whole study population (n=335,689), according to the percentiles of each PRS. Number of participants are shown as n (col%, row%, cell%). Higher percentiles of PRS indicate increased risk of dementia; “≥ 99%” percentile corresponds to the top 1% risk.

| Percentiles of PRS A | Percentiles of PRS B | | | | | | |
| --- | --- | --- | --- | --- | --- | --- | --- |
|  | < 1% | 1-20% | 20-40% | 40-60% | 60-80% | 80-99% | ≥ 99% |
| < 1% | 463  (13.8,  13.8,  0.1) | 2021  (60.2,  3.2,  0.6) | 524  (15.6,  0.8,  0.2) | 215  (6.4,  0.3,  0.1) | 103  (3.1,  0.2,  0.0) | 31  (0.9,  0.0,  0.0) | 0  (0.0,  0.0,  0.0) |
| 1-20% | 2062  (3.2,  61.4,  0.6) | 25986  (40.7, 40.7,  7.7) | 16576  (26.0,  24.7,  4.9) | 10235  (16.0,  15.2,  3.0) | 6060  (9.5,  9.0,  1.8) | 2808  (4.4,  4.4,  0.8) | 54  (0.1,  1.6,  0.0) |
| 20-40% | 520  (0.8,  15.5,  0.2) | 16740  (24.9,  26.2,  5.0) | 17262  (25.7,  25.7,  5.1) | 14719  (21.9,  21.9,  4.4) | 11201  (16.7,  16.7,  3.3) | 6530  (9.7,  10.2,  1.9) | 166  (0.2,  4.9,  0.0) |
| 40-60% | 208  (0.3,  6.2,  0.1) | 10316  (15.4,  16.2,  3.1) | 14802  (22.0,  22.0,  4.4) | 15744  (23.5,  23.5,  4.7) | 14861  (22.1,  22.1,  4.4) | 10814  (16.1,  17.0,  3.2) | 393  (0.6,  11.7,  0.1) |
| 60-80% | 84  (0.1,  2.5,  0.0) | 6100  (9.1,  9.6,  1.8) | 11330  (16.9,  16.9,  3.4) | 14917  (22.2,  22.2,  4.4) | 17482  (26.0,  26.0,  5.2) | 16508  (24.6,  25.9,  4.9) | 716  (1.1,  21.3,  0.2) |
| 80-99% | 20  (0.0,  0.6,  0.0) | 2585  (4.1,  4.1,  0.8) | 6521  (10.2,  9.7,  1.9) | 11008  (17.3,  16.4,  3.3) | 16762  (26.3,  25.0,  5.0) | 25145  (39.4,  39.4,  7.5) | 1740  (2.7,  51.8,  0.5) |
| ≥ 99% | 0  (0.0,  0.0, 0.0) | 33  (1.0,  0.1,  0.0) | 124  (3.7,  0.2,  0.0) | 298  (8.9,  0.4,  0.1) | 669  (19.9,  1.0,  0.2) | 1946  (58.0,  3.1,  0.6) | 287  (8.5, 8.6,  0.1) |

Supplementary Table 5: Cross-classification of predicted risk of breast cancer among participants who experienced the outcome, according to the percentiles of each PRS. Initial score is [PRS-A](https://www.ncbi.nlm.nih.gov/pmc/articles/PMC6323553/), updated score is [PRS-B](https://www.sciencedirect.com/science/article/pii/S0002929720303207?via%3Dihub). 6,347 of 171,490 individuals (3.7%) experienced the outcome. Row %s shown, and may not add up to 100 due to rounding. Categorical NRI (95% CI): 0.013 (-0.006, 0.032).

| Percentiles  of PRS A | Percentiles of PRS B | | | | | | | % reclassified |
| --- | --- | --- | --- | --- | --- | --- | --- | --- |
|  | < 1% | 1-20% | 20-40% | 40-60% | 60-80% | 80-99% | ≥ 99% |  |
| < 1% | 4  (16.0) | 17  (68.0) | 2  (8.0) | 2  (8.0) | 0  (0.0) | 0  (0.0) | 0  (0.0) | 84.0% |
| 1-20% | 8  (1.3) | 217  (36.5) | 186  (31.3) | 106  (17.8) | 62  (10.4) | 16  (2.7) | 0  (0.0) | 63.5% |
| 20-40% | 1  (0.1) | 173  (20.2) | 222  (25.9) | 222  (25.9) | 169  (19.7) | 71  (8.3) | 0  (0.0) | 74.1% |
| 40-60% | 1  (0.1) | 96  (8.4) | 221 (19.4) | 310  (27.3) | 288  (25.3) | 219  (19.3) | 2  (0.2) | 72.7% |
| 60-80% | 0  (0.0) | 56  (3.7) | 192  (12.8) | 278  (18.5) | 445  (29.7) | 511  (34.1) | 18  (1.2) | 70.3% |
| 80-99% | 0  (0.0) | 17  (0.8) | 80  (3.9) | 235  (11.4) | 459  (22.3) | 1132  (55.1) | 131  (6.4) | 44.9% |
| ≥ 99% | 0  (0.0) | 1  (0.6) | 0  (0.0) | 2  (1.1) | 10  (5.6) | 111  (62.4) | 54  (30.3) | 69.7% |

Supplementary Table 6: Cross-classification of predicted risk of hypertension among participants who experienced the outcome, according to the percentiles of each PRS. Initial score is [PRS-A](https://www.nature.com/articles/ng.3768), updated score is [PRS-B](https://www.ncbi.nlm.nih.gov/pmc/articles/PMC6284793/). 137,649 of 317,581 individuals (43.3%) experienced the outcome. Row %s shown, and may not add up to 100 due to rounding. Categorical NRI (95% CI): 0.061 (0.056, 0.067).

| Percentiles of PRS A | Percentiles of PRS B | | | | | | | % reclassified |
| --- | --- | --- | --- | --- | --- | --- | --- | --- |
|  | < 1% | 1-20% | 20-40% | 40-60% | 60-80% | 80-99% | ≥ 99% |  |
| < 1% | 179  (20.4) | 564  (64.3) | 100  (11.4) | 27  (3.1) | 6  (0.7) | 1  (0.1) | 0  (0.0) | 79.6% |
| 1-20% | 496  (2.3) | 9456  (43.5) | 6083  (28.0) | 3556  (16.4) | 1689  (7.8) | 458  (2.1) | 1  (0.0) | 56.5% |
| 20-40% | 71  (0.3) | 5399  (21.3) | 7224  (28.4) | 6230  (24.5) | 4570  (18.0) | 1895  (7.5) | 5  (0.0) | 71.6% |
| 40-60% | 15  (0.1) | 2972  (10.7) | 6049  (21.8) | 7245  (26.1) | 7109  (25.6) | 4305  (15.5) | 45  (0.2) | 73.9% |
| 60-80% | 3  (0.0) | 1292  (4.4) | 3913  (13.4) | 6541  (22.3) | 8917  (30.5) | 8445  (28.8) | 170  (0.6) | 69.5% |
| 80-99% | 2  (0.0) | 355  (1.2) | 1490  (4.8) | 3885  (12.6) | 7883  (25.6) | 15959  (51.8) | 1251  (4.1) | 48.2% |
| ≥ 99% | 0  (0.0) | 1  (0.1) | 6  (0.3) | 38  (2.1) | 142  (7.9) | 1161  (64.8) | 445  (24.8) | 75.2% |

Supplementary Table 7: Cross-classification of predicted risk of dementia among participants who experienced the outcome, according to the percentiles of each PRS. Initial score is [PRS-A](https://doi.org/10.1038/s41467-020-18534-1), updated score is [PRS-B](https://www.ncbi.nlm.nih.gov/pmc/articles/PMC8438688/). 4,460 of 335,689 individuals (1.3%) experienced the outcome. Row %s shown, and may not add up to 100 due to rounding. Categorical NRI (95% CI): 0.057 (0.031, 0.081).

| Percentiles of PRS A | Percentiles of PRS B | | | | | | | % reclassified |
| --- | --- | --- | --- | --- | --- | --- | --- | --- |
|  | < 1% | 1-20% | 20-40% | 40-60% | 60-80% | 80-99% | ≥ 99% |  |
| < 1% | 6  (18.8) | 19  (59.4) | 6  (18.8) | 1  (3.1) | 0  (0.0) | 0  (0.0) | 0  (0.0) | 81.2% |
| 1-20% | 15  (2.1) | 266  (38.1) | 186  (26.6) | 119  (17.0) | 66  (9.5) | 43  (6.2) | 3  (0.4) | 61.9% |
| 20-40% | 7  (0.8) | 165  (20.0) | 213  (25.8) | 172  (20.8) | 151  (18.3) | 116  (14.0) | 3  (0.4) | 74.2% |
| 40-60% | 1  (0.1) | 93  (10.8) | 158  (18.3) | 191  (22.1) | 234  (27.1) | 173  (20.0) | 14  (1.6) | 77.9% |
| 60-80% | 0  (0.0) | 55  (5.9) | 144  (15.5) | 191  (20.5) | 243  (26.1) | 281  (30.2) | 18  (1.9) | 73.9% |
| 80-99% | 0  (0.0) | 22  (2.1) | 90  (8.7) | 170  (16.4) | 235  (22.7) | 468  (45.3) | 49  (4.7) | 54.7% |
| ≥ 99% | 0  (0.0) | 0  (0.0) | 2  (2.7) | 4  (5.5) | 20  (27.4) | 41  (56.2) | 6  (8.2) | 91.8% |

### 5 Tables for top 5% risk

Supplementary Table 8: Cross-classification of predicted risk of breast cancer, according to the percentiles of each PRS. Number of participants are shown as n (col%, row%, cell%). Higher percentiles of PRS indicate increased risk of breast cancer; “≥ 99%” percentile corresponds to the top 1% risk.

| Percentiles  of PRS A | Percentiles of PRS B | | | | | | |
| --- | --- | --- | --- | --- | --- | --- | --- |
|  | < 5% | 5-20% | 20-40% | 40-60% | 60-80% | 80-95% | ≥ 95% |
| < 5% | 2901  (33.8,  33.8,  1.7) | 3319  (38.7,  12.9,  1.9) | 1588  (18.5,  4.6,  0.9) | 582  (6.8,  1.7,  0.3) | 161  (1.9,  0.5,  0.1) | 24  (0.3,  0.1,  0.0) | 1  (0.0,  0.0,  0.0) |
| 5-20% | 3302  (12.8,  38.5,  1.9) | 8489  (33.0,  33.0,  5.0) | 7406  (28.8,  21.6,  4.3) | 4158  (16.2,  12.1,  2.4) | 1900  (7.4,  5.5,  1.1) | 451  (1.8,  1.8,  0.3) | 16  (0.1,  0.2,  0.0) |
| 20-40% | 1569  (4.6,  18.3,  0.9) | 7417  (21.6,  28.8,  4.3) | 9989  (29.1,  29.1,  5.8) | 8053  (23.5,  23.5,  4.7) | 5210  (15.2,  15.2,  3.0) | 1877  (5.5,  7.3,  1.1) | 183  (0.5,  2.1,  0.1) |
| 40-60% | 603  (1.8,  7.0,  0.4) | 4088  (11.9,  15.9,  2.4) | 7998  (23.3,  23.3,  4.7) | 8908  (26.0,  26.0, 5 .2) | 8050  (23.5,  23.5,  4.7) | 4144  (12.1,  16.1,  2.4) | 507  (1.5,  5.9,  0.3) |
| 60-80% | 171  (0.5,  2.0,  0.1) | 1938  (5.7,  7.5,  1.1) | 5296  (15.4,  15.4,  3.1) | 7988  (23.3,  23.3,  4.7) | 10047  (29.3,  29.3,  5.9) | 7400  (21.6,  28.8,  4.3) | 1458  (4.3,  17.0,  0.9) |
| 80-95% | 29  (0.1,  0.3,  0.0) | 453  (1.8,  1.8,  0.3) | 1869  (7.3,  5.4,  1.1) | 4070  (15.8,  11.9,  2.4) | 7402  (28.8,  21.6,  4.3) | 8554  (33.3,  33.3,  5.0) | 3346  (13.0,  39.0,  2.0) |
| ≥ 95% | 0  (0.0,  0.0,  0.0) | 19  (0.2,  0.1,  0.0) | 152  (1.8,  0.4,  0.1) | 539  (6.3,  1.6,  0.3) | 1528  (17.8,  4.5,  0.9) | 3273  (38.2,  12.7,  1.9) | 3064  (35.7,  35.7,  1.8) |

Supplementary Table 9: Cross-classification of predicted risk of hypertension, according to the percentiles of each PRS. Number of participants are shown as n (col%, row%, cell%). Higher percentiles of PRS indicate increased risk of hypertension; “≥ 99%” percentile corresponds to the top 1% risk.

| Percentiles of PRS A | Percentiles of PRS B | | | | | | |
| --- | --- | --- | --- | --- | --- | --- | --- |
|  | < 5% | 5-20% | 20-40% | 40-60% | 60-80% | 80-95% | ≥ 95% |
| < 5% | 5721  (36.0,  36.0,  1.8) | 6110  (38.5,  12.8,  1.9) | 2784  (17.5,  4.4,  0.9) | 940  (5.9,  1.5,  0.3) | 281  (1.8,  0.4,  0.1) | 42  (0.3,  0.1,  0.0) | 2  (0.0,  0.0,  0.0) |
| 5-20% | 6156  (12.9,  38.8,  1.9) | 16143  (33.9,  33.9,  5.1) | 13499  (28.3,  21.3,  4.3) | 7569  (15.9,  11.9,  2.4) | 3388  (7.1,  5.3,  1.1) | 830  (1.7,  1.7,  0.3) | 52  (0.1,  0.3,  0.0) |
| 20-40% | 2772  (4.4,  17.5,  0.9) | 13532  (21.3,  28.4,  4.3) | 18776  (29.6,  29.6,  5.9) | 14854  (23.4,  23.4,  4.7) | 9954  (15.7,  15.7,  3.1) | 3375  (5.3,  7.1,  1.1) | 253  (0.4,  1.6,  0.1) |
| 40-60% | 943  (1.5,  5.9,  0.3) | 7516  (11.8,  15.8,  2.4) | 14996  (23.6,  23.6,  4.7) | 16595  (26.1,  26.1,  5.2) | 15006  (23.6,  23.6,  4.7) | 7475  (11.8,  15.7,  2.4) | 985  (1.6,  6.2,  0.3) |
| 60-80% | 248  (0.4,  1.6,  0.1) | 3430  (5.4,  7.2,  1.1) | 9803  (15.4,  15.4,  3.1) | 14988  (23.6,  23.6,  4.7) | 18520  (29.2,  29.2,  5.8) | 13721  (21.6,  28.8,  4.3) | 2806  (4.4,  17.7,  0.9) |
| 80-95% | 40  (0.1,  0.3,  0.0) | 839  (1.8,  1.8,  0.3) | 3390  (7.1,  5.3,  1.1) | 7640  (16.0,  12.0,  2.4) | 13584  (28.5,  21.4,  4.3) | 16051  (33.7,  33.7,  5.1) | 6093  (12.8,  38.4,  1.9) |
| ≥ 95% | 0  (0.0,  0.0,  0.0) | 67  (0.4,  0.1,  0.0) | 270  (1.7,  0.4,  0.1) | 929  (5.9,  1.5,  0.3) | 2782  (17.5,  4.4,  0.9) | 6143  (38.7,  12.9,  1.9) | 5688  (35.8,  35.8,  1.8) |

Supplementary Table 10: Cross-classification of predicted risk of dementia, according to the percentiles of each PRS. Number of participants are shown as n (col%, row%, cell%). Higher percentiles of PRS indicate increased risk of dementia; “≥ 99%” percentile corresponds to the top 1% risk.

| Percentiles of PRS A | Percentiles of PRS B | | | | | | |
| --- | --- | --- | --- | --- | --- | --- | --- |
|  | < 5% | 5-20% | 20-40% | 40-60% | 60-80% | 80-95% | ≥ 95% |
| < 5% | 4500  (26.8,  26.8,  1.3) | 5846  (34.8,  11.6,  1.7) | 3547  (21.1,  5.3,  1.1) | 1718  (10.2,  2.6,  0.5) | 838  (5.0,  1.2,  0.2) | 295  (1.8,  0.6,  0.1) | 41  (0.2,  0.2,  0.0) |
| 5-20% | 5896  (11.7,  35.1,  1.8) | 14290  (28.4,  28.4,  4.3) | 13553  (26.9,  20.2,  4.0) | 8732  (17.3,  13.0, 2.6) | 5325  (10.6,  7.9,  1.6) | 2169  (4.3,  4.3,  0.6) | 388  (0.8,  2.3,  0.1) |
| 20-40% | 3597  (5.4,  21.4,  1.1) | 13663  (20.4,  27.1,  4.1) | 17262  (25.7,  25.7,  5.1) | 14719  (21.9,  21.9,  4.4) | 11201  (16.7,  16.7,  3.3) | 5649  (8.4,  11.2,  1.7) | 1047  (1.6,  6.2,  0.3) |
| 40-60% | 1727  (2.6,  10.3,  0.5) | 8797  (13.1,  17.5,  2.6) | 14802  (22.0,  22.0,  4.4) | 15744  (23.5,  23.5,  4.7) | 14861  (22.1,  22.1,  4.4) | 9026  (13.4,  17.9, 2.7) | 2181  (3.2,  13.0,  0.6) |
| 60-80% | 802  (1.2,  4.8,  0.2) | 5382  (8.0,  10.7,  1.6) | 11330  (16.9,  16.9,  3.4) | 14917  (22.2,  22.2,  4.4) | 17482  (26.0,  26.0,  5.2) | 13340  (19.9,  26.5,  4.0) | 3884  (5.8,  23.1,  1.2) |
| 80-95% | 235  (0.5,  1.4,  0.1) | 2110  (4.2, 4.2,  0.6) | 5638  (11.2,  8.4,  1.7) | 9254  (18.4,  13.8,  2.8) | 13497  (26.8,  20.1,  4.0) | 14050  (27.9,  27.9,  4.2) | 5569  (11.1,  33.2, 1.7) |
| ≥ 95% | 28  (0.2,  0.2,  0.0) | 265  (1.6,  0.5,  0.1) | 1007  (6.0,  1.5,  0.3) | 2052  (12.2,  3.1,  0.6) | 3934  (23.4, 5.9,  1.2) | 5824  (34.7,  11.6,  1.7) | 3675 (21.9,  21.9,  1.1) |

Supplementary Table 11: Cross-classification of predicted risk of breast cancer among participants who experienced the outcome, according to the percentiles of each PRS. Initial score is [PRS-A](https://www.ncbi.nlm.nih.gov/pmc/articles/PMC6323553/), updated score is PRS-B. 6,347 of 171,490 individuals (3.7%) experienced the outcome. Row %s shown, and may not add up to 100 due to rounding. Categorical NRI (95% CI): 0.011 (-0.010, 0.030).

| Percentiles of PRS A | Percentiles of PRS B | | | | | | | % reclassified |
| --- | --- | --- | --- | --- | --- | --- | --- | --- |
|  | < 5% | 5-20% | 20-40% | 40-60% | 60-80% | 80-95% | ≥ 95% |  |
| < 5% | 31  (29.0) | 41  (38.3) | 23  (21.5) | 10  (9.3) | 2  (1.9) | 0  (0.0) | 0  (0.0) | 71.0% |
| 5-20% | 30  (5.8) | 144  (28.1) | 165  (32.2) | 98  (19.1) | 60  (11.7) | 16  (3.1) | 0  (0.0) | 71.9% |
| 20-40% | 16  (1.9) | 158  (18.4) | 222  (25.9) | 222  (25.9) | 169  (19.7) | 66  (7.7) | 5  (0.6) | 74.1% |
| 40-60% | 12  (1.1) | 85  (7.5) | 221  (19.4) | 310  (27.3) | 288  (25.3) | 188  (16.5) | 33  (2.9) | 72.7% |
| 60-80% | 3  (0.2) | 53  (3.5) | 192  (12.8) | 278  (18.5) | 445  (29.7) | 426  (28.4) | 103  (6.9) | 70.3% |
| 80-95% | 1  (0.1) | 15  (1.0) | 74  (4.9) | 213  (14.0) | 383  (25.2) | 568  (37.3) | 267  (17.6) | 62.7% |
| ≥ 95% | 0  (0.0) | 2  (0.3) | 6  (0.8) | 24  (3.4) | 86  (12.1) | 257  (36.1) | 336  (47.3) | 52.7% |

Supplementary Table 12: Cross-classification of predicted risk of hypertension among participants who experienced the outcome, according to the percentiles of each PRS. Initial score is [PRS-A](https://www.nature.com/articles/ng.3768), updated score is [PRS-B](https://www.ncbi.nlm.nih.gov/pmc/articles/PMC6284793/). 137,649 of 317,581 individuals (43.3%) experienced the outcome. Row %s shown, and may not add up to 100 due to rounding. Categorical NRI (95% CI): 0.064 (0.058, 0.070).

| Percentiles of PRS A | Percentiles of PRS B | | | | | | | % reclassified |
| --- | --- | --- | --- | --- | --- | --- | --- | --- |
|  | < 5% | 5-20% | 20-40% | 40-60% | 60-80% | 80-95% | ≥ 95% |  |
| < 5% | 1509  (30.2) | 1940  (38.8) | 1031  (20.6) | 367  (7.3) | 127  (2.5) | 21  (0.4) | 1  (0.0) | 69.8% |
| 5-20% | 1757  (10.0) | 5489  (31.2) | 5152  (29.2) | 3216  (18.3) | 1568  (8.9) | 416  (2.4) | 22  (0.1) | 68.8% |
| 20-40% | 825  (3.2) | 4645  (18.3) | 7224  (28.4) | 6230  (24.5) | 4570  (18.0) | 1762  (6.9) | 138  (0.5) | 71.6% |
| 40-60% | 274  (1.0) | 2713  (9.8) | 6049  (21.8) | 7245  (26.1) | 7109  (25.6) | 3803  (13.7) | 547  (2.0) | 73.9% |
| 60-80% | 86  (0.3) | 1209  (4.1) | 3913  (13.4) | 6541  (22.3) | 8917  (30.5) | 7067  (24.1) | 1548  (5.3) | 69.5% |
| 80-95% | 17  (0.1) | 312  (1.3) | 1381  (5.8) | 3478  (14.5) | 6648  (27.8) | 8541  (35.7) | 3538  (14.8) | 64.3% |
| ≥ 95% | 0  (0.0) | 29  (0.3) | 115  (1.3) | 445  (5.1) | 1377  (15.8) | 3358  (38.6) | 3379  (38.8) | 61.2% |

Supplementary Table 13: Cross-classification of predicted risk of dementia among participants who experienced the outcome, according to the percentiles of each PRS. Initial score is [PRS-A](https://doi.org/10.1038/s41467-020-18534-1), updated score is [PRS-B](https://www.ncbi.nlm.nih.gov/pmc/articles/PMC8438688/). 4,460 of 335,689 individuals (1.3%) experienced the outcome. Row %s shown, and may not add up to 100 due to rounding. Categorical NRI (95% CI): 0.06 (0.033, 0.085).

| Percentiles of PRS A | Percentiles of PRS B | | | | | | | % reclassified |
| --- | --- | --- | --- | --- | --- | --- | --- | --- |
|  | < 5% | 5-20% | 20-40% | 40-60% | 60-80% | 80-95% | ≥ 95% |  |
| < 5% | 45  (28.0) | 61  (37.9) | 38  (23.6) | 11  (6.8) | 1  (0.6) | 3  (1.9) | 2  (1.2) | 72.0% |
| 5-20% | 50  (8.8) | 150  (26.4) | 154  (27.1) | 109  (19.2) | 65  (11.4) | 34  (6.0) | 7  (1.2) | 73.6% |
| 20-40% | 30  (3.6) | 142  (17.2) | 213  (25.8) | 172  (20.8) | 151  (18.3) | 102  (12.3) | 17  (2.1) | 74.2% |
| 40-60% | 12  (1.4) | 82  (9.5) | 158  (18.3) | 191  (22.1) | 234  (27.1) | 147  (17.0) | 40  (4.6) | 77.9% |
| 60-80% | 6  (0.6) | 49  (5.3) | 144  (15.5) | 191  (20.5) | 243  (26.1) | 226  (24.2) | 73  (7.8) | 73.9% |
| 80-95% | 0  (0.0) | 19  (2.4) | 81  (10.3) | 132  (16.8) | 185  (23.5) | 249  (31.6) | 122  (15.5) | 68.4% |
| ≥ 95% | 45  (28.0) | 61  (37.9) | 38  (23.6) | 11  (6.8) | 1  (0.6) | 3  (1.9) | 2  (1.2) | 72.0% |

### 6 ICD codes for disease ascertainment

Supplementary Table 14: ICD and OPCS codes for breast cancer outcome and exclusions. Individuals with prior breast cancer, carcinoma in situ or mastectomy were excluded. Breast cancer outcomes were identified from cancer registry, hospital episode statistics and death registry.

| Breast cancer | | Breast carcinoma in situ | Mastectomy |
| --- | --- | --- | --- |
| ICD-10 | **ICD-9** | **ICD-10** | **OPCS-4** |
| C50.0 | 1740 | D05.0 | B27.1 |
| C50.1 | 1741 | D05.1 | B27.2 |
| C50.2 | 1742 | D05.7 | B27.3 |
| C50.3 | 1743 | D05.9 | B27.4 |
| C50.4 | 1744 |  | B27.5 |
| C50.5 | 1745 |  | B27.6 |
| C50.6 | 1746 |  | B27.8 |
| C50.8 | 1748 |  | B27.9 |
| C50.9 | 1749 |  | B28.1 |
|  |  |  | B28.2 |
|  |  |  | B28.3 |
|  |  |  | B28.4 |
|  |  |  | B28.6 |

Supplementary Table 15: ICD codes for hypertension exclusions. Individuals with prior major adverse cardiovascular events (MACE) were excluded; MACE was identified from hospital episode statistics and UK Biobank baseline interview and touchscreen questionnaire answers (UK Biobank Data Fields [20002](https://biobank.ndph.ox.ac.uk/showcase/field.cgi?id=20002), [20004](https://biobank.ndph.ox.ac.uk/showcase/field.cgi?id=20004) and [6150](https://biobank.ndph.ox.ac.uk/showcase/field.cgi?id=6150))

| **MACE** | | | | |
| --- | --- | --- | --- | --- |
| **Hospital Episode Statistics** | | | **Verbal Interview** | |
| **ICD-10** | **ICD-9** | **OPCS-4** | **Non-cancer illness** | **Operation** |
| I20.0 | 4109 | K40.1 | 1075 | 1070 |
| I21.0 | 4119 | K40.2 | 1081 | 1095 |
| I21.1 | 4129 | K40.3 | 1082 | 1105 |
| I21.2 | 4309 | K40.4 | 1086 | 1109 |
| I21.3 | 4319 | K40.8 | 1491 | 1523 |
| I21.4 | 4330 | K40.9 | 1583 |  |
| I21.9 | 4331 | K41.1 |  |  |
| I21.X | 4332 | K41.2 |  |  |
| I22.0 | 4333 | K41.3 |  |  |
| I22.1 | 4338 | K41.4 |  |  |
| I22.8 | 4339 | K41.8 |  |  |
| I22.9 | 4340 | K41.9 |  |  |
| I23.0 | 4341 | K42.1 |  |  |
| I23.1 | 4349 | K42.2 |  |  |
| I23.2 | 4359 | K42.3 |  |  |
| I23.3 | 4369 | K42.4 |  |  |
| I23.4 |  | K42.8 |  |  |
| I23.5 |  | K42.9 |  |  |
| I23.6 |  | K43.1 |  |  |
| I23.8 |  | K43.2 |  |  |
| I25.2 |  | K43.3 |  |  |
| I25.6 |  | K43.4 |  |  |
| I60.0 |  | K43.8 |  |  |
| I60.1 |  | K43.9 |  |  |
| I60.2 |  | K44.1 |  |  |
| I60.3 |  | K44.2 |  |  |
| I60.4 |  | K44.8 |  |  |
| I60.5 |  | K44.9 |  |  |
| I60.6 |  | K45.1 |  |  |
| I60.7 |  | K45.2 |  |  |
| I60.8 |  | K45.3 |  |  |
| I60.9 |  | K45.4 |  |  |
| I61.0 |  | K45.5 |  |  |
| I61.1 |  | K45.6 |  |  |
| I61.2 |  | K45.8 |  |  |
| I61.3 |  | K45.9 |  |  |
| I61.4 |  | K46.1 |  |  |
| I61.5 |  | K46.2 |  |  |
| I61.6 |  | K46.3 |  |  |
| I61.8 |  | K46.4 |  |  |
| I61.9 |  | K46.5 |  |  |
| I63.0 |  | K46.8 |  |  |
| I63.1 |  | K46.9 |  |  |
| I63.2 |  | K47.1 |  |  |
| I63.3 |  | K48.3 |  |  |
| I63.4 |  | K49.1 |  |  |
| I63.5 |  | K49.2 |  |  |
| I63.6 |  | K49.3 |  |  |
| I63.8 |  | K49.4 |  |  |
| I63.9 |  | K49.8 |  |  |
| I64.4 |  | K49.9 |  |  |
| G45.0 |  | K50.1 |  |  |
| G45.1 |  | K50.2 |  |  |
| G45.2 |  | K50.3 |  |  |
| G45.3 |  | K50.4 |  |  |
| G45.4 |  | K50.8 |  |  |
| G45.8 |  | K50.9 |  |  |
| G45.9 |  | K75.1 |  |  |
| G46.0 |  | K75.2 |  |  |
| G46.1 |  | K75.3 |  |  |
| G46.2 |  | K75.4 |  |  |
| G46.3 |  | K75.8 |  |  |
| G46.4 |  | K75.9 |  |  |
| G46.5 |  | L29.1 |  |  |
| G46.6 |  | L29.2 |  |  |
| G46.7 |  | L29.3 |  |  |
| G46.8 |  | L29.4 |  |  |
|  |  | L29.5 |  |  |
|  |  | L29.6 |  |  |
|  |  | L29.7 |  |  |
|  |  | L30.3 |  |  |
|  |  | L31.1 |  |  |
|  |  | L31.4 |  |  |
|  |  | L34.3 |  |  |
|  |  | L35.3 |  |  |
|  |  | L35.4 |  |  |

Supplementary Table 16: ICD codes for Dementia outcome and exclusions. Individuals with prior dementia were excluded. Dementia/Alzheimer’s disease outcomes were identified from hospital episode statistics and death registry.

| Dementia | |
| --- | --- |
| ICD-10 | **ICD-9** |
| A81.0 | 2902 |
| I67.3 | 2903 |
| F00.0 | 2904 |
| F00.1 | 2912 |
| F00.2 | 2941 |
| F00.9 | 3310 |
| F01.0 | 3311 |
| F01.1 | 3312 |
| F01.2 | 3315 |
| F01.3 |  |
| F01.8 |  |
| F01.9 |  |
| F02.0 |  |
| F02.1 |  |
| F02.2 |  |
| F02.3 |  |
| F02.4 |  |
| F02.8 |  |
| F03 |  |
| F05.1 |  |
| F10.6 |  |
| G30.0 |  |
| G30.1 |  |
| G30.8 |  |
| G30.9 |  |
| G31.1 |  |
| G31.8 |  |
